# Changing characteristics over time of individuals receiving COVID-19 vaccines in Denmark: A population-based descriptive study of vaccine uptake

**DOI:** 10.1101/2022.03.25.22272927

**Authors:** Mette Reilev, Morten Olesen, Helene Kildegaard, Henrik Støvring, Jacob H. Andersen, Jesper Hallas, Lars C. Lund, Louise Ladebo, Martin T. Ernst, Per Damkier, Peter B. Jensen, Anton Pottegård, Lotte Rasmussen

## Abstract

**Aims:** The Danish authorities implemented a differential rollout of the COVID-19 vaccines where individuals at high risk of COVID-19 were prioritized. We describe the temporal uptake of COVID-19 vaccines and changing characteristics of vaccine recipients in Denmark.

**Methods:** Using the Danish national health care registries, we identified all Danish residents ≥5 years of age who received at least one dose of a COVID-19 vaccine from December 27^th^, 2020, to January 29^th^, 2022. We charted the daily number of newly vaccinated individuals and the cumulative vaccine coverage over time, stratified by vaccine type, age groups, and vaccination priority groups. In addition, we described characteristics of vaccine recipients during 2-months-intervals and in vaccination priority groups.

**Results:** By January 29^th^, 2022, 86%, 84% and 63% of Danish residents ≥5 years had received a first, second, and third dose, respectively, of a COVID-19 vaccine, most commonly the BNT162b2 vaccine (84% of vaccinated individuals). Vaccine uptake ranged from 48% in 5-11-year-olds up to 98% in 65-74-year-olds. Individuals vaccinated before June 2021 were older (median age 61-70 years vs. 10-35 years in later periods) and had more comorbidities such as hypertension (22-28% vs. 0.77-2.8% in later periods), chronic lung disease (9.4-15% vs. 3.7-4.6% in later periods), and diabetes (9.3-12% vs. 0.91-2.4% in later periods).

**Conclusions:** The uptake of the COVID-19 vaccines is high in Denmark. We document substantial changes over time in characteristics of vaccine recipients which should be considered when designing and interpreting studies on the effectiveness and safety of COVID-19 vaccines.

## Background

The initially limited supply capacity and high demand for coronavirus disease 2019 (COVID-19) vaccines forced authorities worldwide to implement a differential rollout of vaccines such that individuals at highest risk of severe COVID-19 were prioritized before younger and healthier individuals. To inform the design and interpretation of real-world studies on both the effectiveness and safety of the COVID-19 vaccines as well as on the course of the COVID-19 pandemic, we described the uptake of the COVID-19 vaccines in Denmark including characteristics of individuals receiving the COVID-19 vaccines at different time periods.

## Methods

### Data sources

From the Danish Vaccination registry^1^ we identified all Danish residents ≥5 years of age at the time of receiving the first dose of one of the four COVID-19 vaccines used in Denmark: BNT162b2 (Comirnaty®, Pfizer BioNTech), mRNA-1273 (Spikevax®, Moderna), AZD1222 (Vaxzevria®, AstraZeneca), and Ad26.COV2.S (COVID-19 Vaccine Janssen, Johnson&Johnson). Individual-level data were linked via a unique personal identifier to the Danish National Patient^2^ and Prescription Registry^3^ to obtain information on discharge diagnoses and prescription drug use. Information on positive and negative real time reverse transcription polymerase chain reaction (RT-PCR) tests for severe acute respiratory syndrome coronavirus 2 (SARS-CoV-2) was obtained from the Danish Microbiology Database.^4^

### Vaccine rollout strategy

Denmark began its COVID-19 vaccination program on December 27^th^, 2020 (**Supplementary Figure 1**). The Danish health authorities initially prioritized immunization of nursing home residents, frontline healthcare- and social workers, and persons at particularly high risk of severe COVID-19 (see **Appendix A** for a detailed description of vaccination priority groups). From mid-March to end-November 2021, the vaccination program was gradually extended to the remaining population ≥5 years of age. Danish residents have mainly been offered either the BNT162b2 or mRNA-1273 vaccine. The AZD1222 vaccine was prioritized for healthcare and social workers, but removed from the vaccination program on March 11, 2021, due to safety concerns.^5^ The Ad26.COV2.S vaccine was only available on a voluntary opt-in basis.

### Analysis

We described the daily number of individuals receiving a first vaccine dose against COVID-19 and the cumulative vaccination coverage from December 27^th^, 2020, to January 29^th^, 2022, stratified by vaccine type, age groups, and vaccination priority groups. We described the overall uptake of the first, second, and third dose, and in age groups and vaccination priority groups. Finally, we described characteristics of recipients of a first vaccination dose within two months-intervals. We stratified by type of vaccine, vaccination priority group, whether they received the second dose within 8 weeks, and whether they had a prior positive SARS-CoV-2 PCR test. Codes used to define comorbidity and prior medical history are available in **Supplementary Table 1**.

### Other

According to Danish law, studies based entirely on registry data do not require approval from an ethics review board ^6^. The study was registered at the repository of the University of Southern Denmark(10.960) and data were available from the Danish Health Data Authority(FSEID00005447). Due to legal reasons, individual-level data cannot be shared by the authors.

## Results

By January 29^th^, 2022, 86%, 84%, and 63% of the Danish population ≥5 years of age had received a first, second, and third dose, respectively, of a COVID-19 vaccine (**Data not shown**). Uptake of the first vaccination dose ranged from 48% in 5-11-year-olds up to 98% in 65-74-year-olds (**Supplementary Figure 2**). Most received the BNT162b2 vaccine (n=4,114,960, 84%) (**Supplementary Figure 3**), primarily from March through July 2021 (**Supplementary Figure 4**). The rollout of the first vaccination dose in the highest prioritized groups was largely accomplished before April 2021 (**Figure 1A**) whereas most individuals ≥75 years had received the first dose by the end of May 2021 (**Figure 1B, Supplementary Figure 2**). Overall, 98% and 73% of vaccinated individuals received a second and third dose, respectively, before January 29^th^ (**Supplementary Table 2**). Of these, 97% received homologous first- and second doses, whereas 99% received homologous second- and third doses (**Supplementary Table 3**).

**Figure 1.**
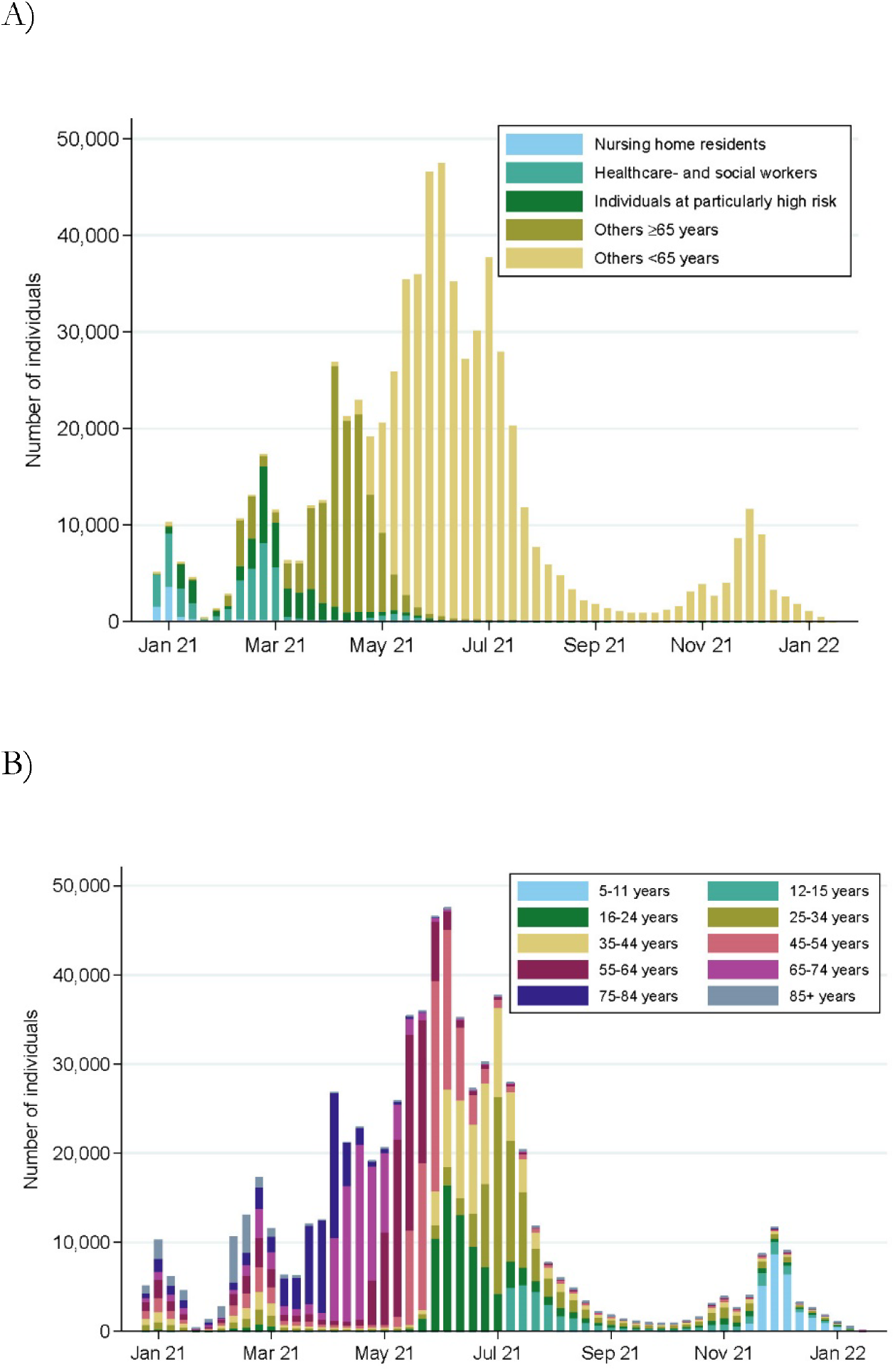
The number of individuals who received their first dose of COVID-19 vaccine each day from December 27^th^, 2020, to January 29^th^, 2022, stratified by A) vaccine priority groups, and B) age groups.

The proportion of women among vaccine recipients changed from 70% in December 2020-January 2021 to ∼50% after April 2021 (**Figure 2**) reflecting a high proportion of women among nursing home residents (62%), and healthcare and social workers (80%) (**Figure 3**). The median age at time of vaccination was 61 years (IQR 46-81) in December 2020-January 2021, increasing to 70 years (IQR 50-82) in February-March 2021, and decreasing to 10 years (IQR 8-13) in December 2021-January 2022 (**Figure 2**). Individuals vaccinated before June 2021 had more comorbidities than in later periods including hypertension (22-28% vs. 0.77-2.8%), chronic lung disease (9.4-15% vs. 3.7-4.6%), and diabetes (9.3-12% vs. 0.91-2.4%) (**Figure 2**). The prevalence of comorbidities among vaccinated individuals generally decreased over time (**Figure 2**). The prevalence and type of comorbidities varied markedly across vaccination priority groups (**Figure 3**).

**Figure 2.**
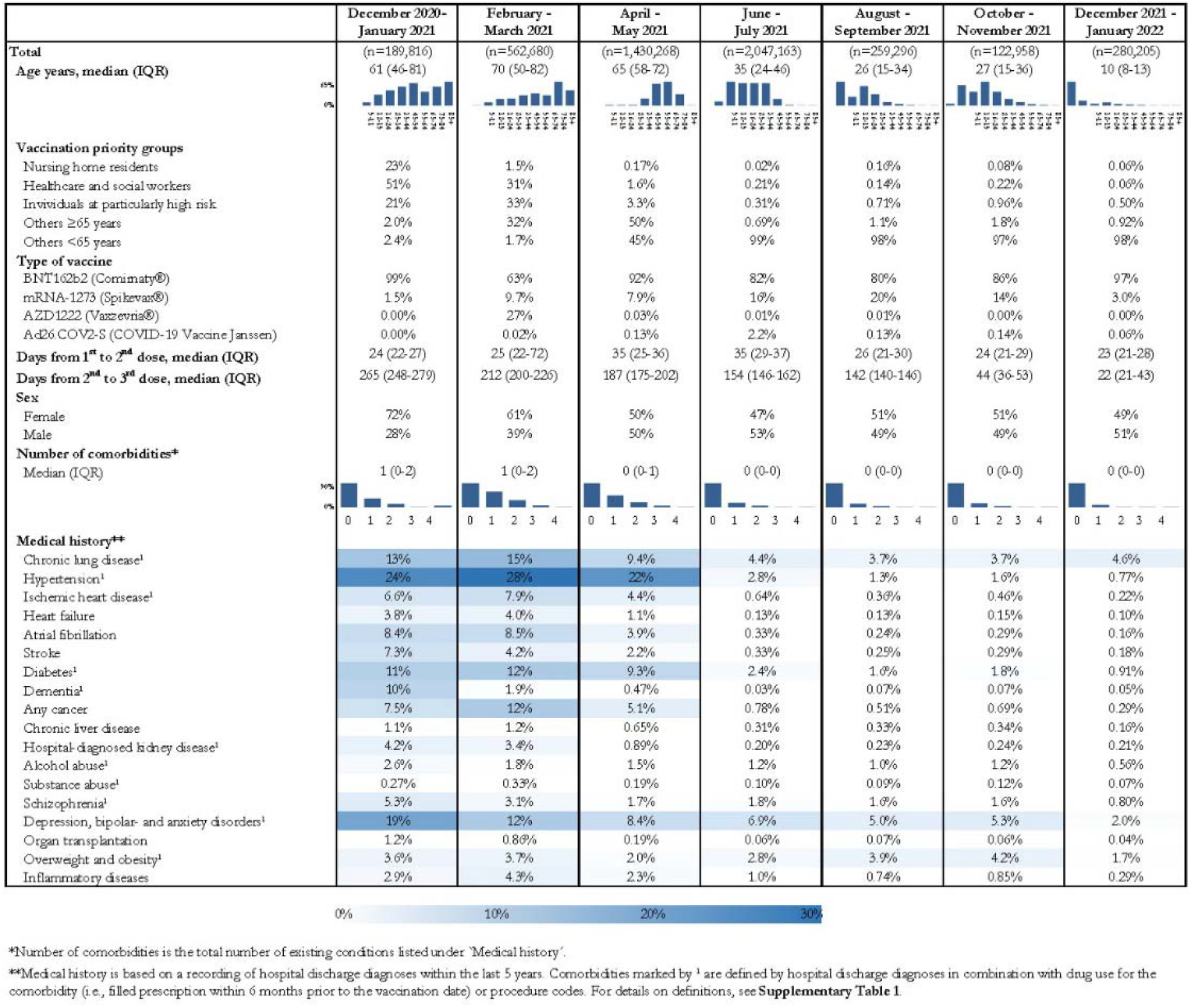
Characteristics within two-months intervals of individuals receiving their first dose of a COVID-19 vaccine.

**Figure 3.**
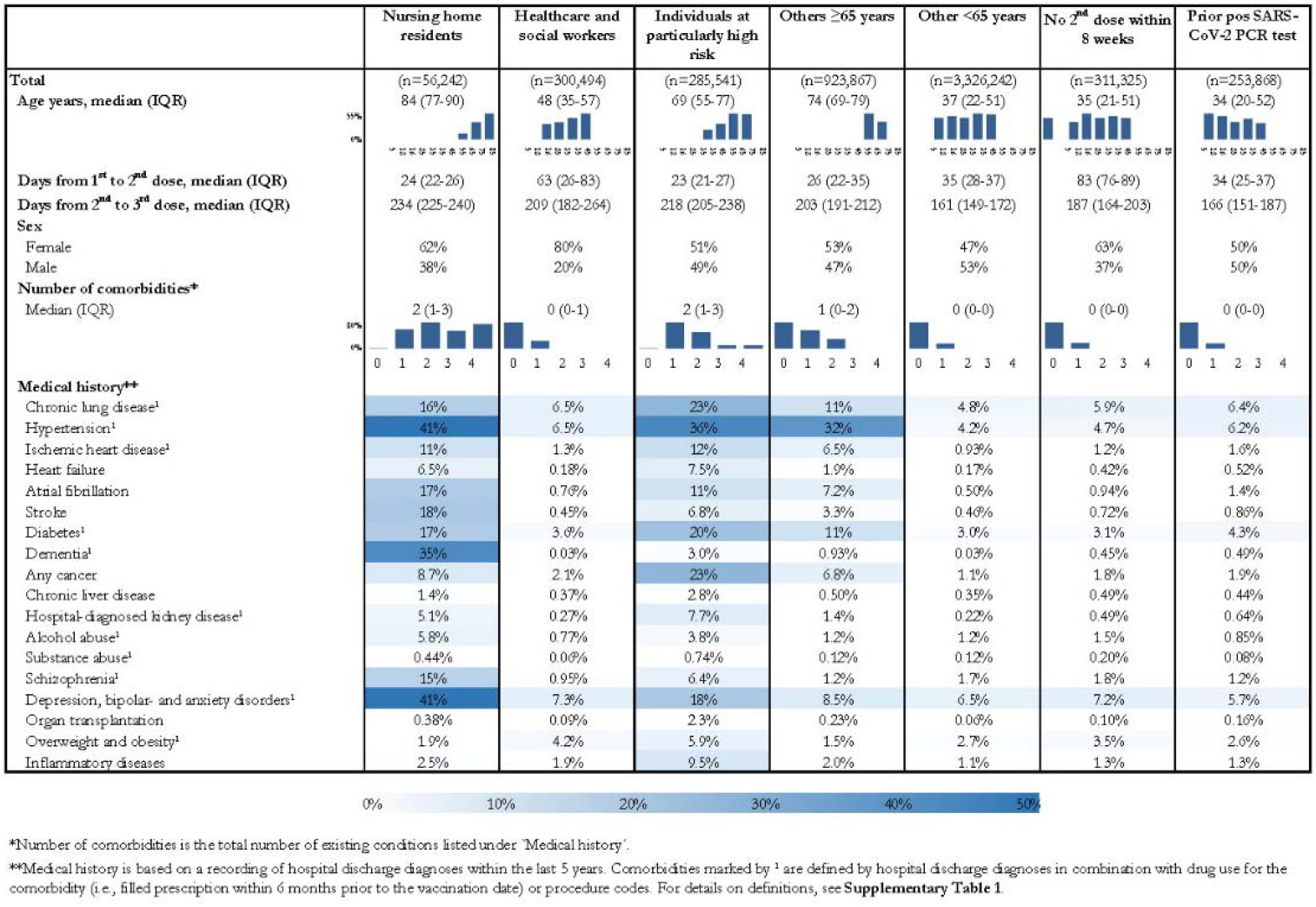
Characteristics of individuals receiving their first dose of a COVID-19 vaccine, stratified by priority group, whether they did not receive their 2^nd^ dose within 8 weeks, and whether they had a positive SARS-CoV-2 PCR test prior to receiving their first vaccination dose.

## Discussion

We have documented an extensive uptake of COVID-19 vaccines in Denmark where 86% of the eligible population had received at least one dose by January 29^th^, 2022. Individuals vaccinated before April 2021 were more often women, people of older age and had more comorbidities compared to those vaccinated in the remaining period, reflecting the differential rollout which initially prioritized individuals at particularly high risk of COVID-19.

The highly valid Danish health registries^2^ enabled a nationwide description of the COVID-19 vaccine uptake including a comprehensive description of medical history among individuals receiving the COVID-19 vaccine at different time points. Though data from clinical databases and on diagnoses from primary care were not available, we have been able to identify the most relevant medical history by use of hospital discharge diagnoses and drug use from primary care. Information on individuals not accepting a COVID-19 vaccine was not available. Also, we did not have access to data on socioeconomic factors such as income, educational level, and migration status which have been shown to be associated with vaccine acceptance in both low- and middle-income^7^ and high-income countries.^8^

Though the observed changes in patient characteristics over time among those vaccinated are specifically related to the vaccine rollout strategy applied in Denmark, similar strategies were implemented in other countries. As such, most countries initially prioritized vaccination of high-risk individuals and health care workers, and later expanded vaccine rollout, primarily vaccinating the oldest before the youngest.^9–12^

The Danish health registries are well-suited for timely analyses of COVID-19 vaccine safety and effectiveness, due to their high validity and frequently updated information on vaccine uptake. However, the analyses and interpretation of such studies should consider the marked differences in patient characteristics over time and across vaccine priority groups. Since important differences in patient characteristics also exist across recipients of different vaccines, similar considerations should be undertaken in head-to-head comparisons of vaccine effectiveness. Importantly, changes over time in characteristics of patients receiving the COVID-19 vaccines should be considered in any study relying on data from countries that implemented a differential rollout strategy. Similar considerations of changes in characteristics also applies for individuals not accepting the vaccine.

## Conclusions

COVID-19 vaccines had a high uptake in Denmark, though characterized by noticeable differences in characteristics over time among vaccine recipients. This should be considered when designing and interpreting studies on the course of the COVID-19 pandemic, and on the effectiveness and safety of COVID-19 vaccines.

## Data Availability

Due to legal reasons, individual level raw data from Danish administrative and health registries cannot be shared by the authors.

## Acknowledgements

None to report.

## References

1. Grove Krause T, Jakobsen S, Haarh M, et al. The Danish vaccination register. Euro Surveill Bull Eur Sur Mal Transm Eur Commun Dis Bull 2012; 17: 20155.

2. Schmidt M, Schmidt SAJ, Sandegaard JL, et al. The Danish National Patient Registry: a review of content, data quality, and research potential. Clin Epidemiol 2015; 449.

3. Pottegård A, Schmidt SAJ, Wallach-Kildemoes H, et al. Data Resource Profile: The Danish National Prescription Registry. Int J Epidemiol 2017; 46: 798–798f.

4. Voldstedlund M, Haarh M, Mølbak K, et al. The Danish Microbiology Database (MiBa) 2010 to 2013. Euro Surveill Bull Eur Sur Mal Transm Eur Commun Dis Bull 2014; 19: 20667.

5. Wise J. Covid-19: European countries suspend use of Oxford-AstraZeneca vaccine after reports of blood clots. BMJ 2021; n699.

6. Thygesen LC, Daasnes C, Thaulow I, et al. Introduction to Danish (nationwide) registers on health and social issues: structure, access, legislation, and archiving. Scand J Public Health 2011; 39: 12–16.

7. Moola S, Gudi N, Nambiar D, et al. A rapid review of evidence on the determinants of and strategies for COVID-19 vaccine acceptance in low- and middle-income countries. J Glob Health 2021; 11: 05027.

8. The New York Times. Tracking Coronavirus Vaccinations Around the World 2021, https://www.nytimes.com/interactive/2021/world/covid-vaccinations-tracker.html (2022, accessed 21 January 2022).

9. European Centre for Disease Prevention and Control. Overview of the implementation of COVID-19 vaccination strategies and deployment plans in the EU/EEA, https://www.ecdc.europa.eu/sites/default/files/documents/COVID-19-vaccination-strategies-and-deployment-plans-Nov-2021.pdf (2021, accessed 21 January 2022).

10. Dooling K, Marin M, Wallace M, et al. The Advisory Committee on Immunization Practices’ Updated Interim Recommendation for Allocation of COVID-19 Vaccine - United States, December 2020. MMWR Morb Mortal Wkly Rep 2021; 69: 1657–1660.

11. Rosen B, Waitzberg R, Israeli A. Israel’s rapid rollout of vaccinations for COVID-19. Isr J Health Policy Res 2021; 10: 6.

12. Australia’s COVID-19 vaccine national roll-out strategy, https://www.health.gov.au/sites/default/files/documents/2021/01/covid-19-vaccination-australia-s-covid-19-vaccine-national-roll-out-strategy.pdf (accessed 21 January 2022).

