## Supplementary for "Changing characteristics over time of individuals receiving COVID-19 vaccines in Denmark: A population-based descriptive study of vaccine uptake"

**Appendix A. Vaccination priority groups**

The Danish COVID-19 vaccination program was rolled out according to twelve priority groups. Individuals in priority group 1-2 and 4-6 were called for vaccination simultaneously when the first COVID-19 vaccines became available from December 27^th^, 2020 and onwards. These groups consisted of nursing home residents (priority group 1), persons ≥65 years of age who receive personal care and practical support at home (priority group 2), frontline healthcare- and social workers (priority group 4), persons over 65 years of age at particular risk of a severe course of COVID-19 (priority group 5) as well as indispensable caregivers for acute and chronically ill patients (priority group 6). Persons ≥65 years of age at particular risk of a severe course of COVID-19 were defined in a letter from the Danish Health Authorities on December 26^th^, 2020 and sent to all Danish hospitals and general practitioners who were asked to identify and refer such persons for vaccination (ref1). Persons at particular risk of severe COVID-19 included persons with a high disease severity of one or more of the following conditions: primary or acquired immunodeficiencies, immunosuppressant therapy, organ transplants, heart failure, ischemic heart disease, hypertension, congenital heart disease, valvular heart disease, chronic obstructive pulmonary disease, infectious lung disease, interstitial lung disease, genetic lung diseases, other conditions with chronic respiratory failure, chronic kidney disease, chronic liver disease, disseminated cancer or ongoing treatment of cancer, diabetes, obesity, cognitive impairment and neurological or rheumatic disorders with cough impairment. Priority group 5 of persons at particularly risk of severe COVID-19 was in March 2021 extended to also include individuals under 65 years of age with these diseases as well as individuals with severe psychiatric disease, children with certain chronic diseases and pregnant women (ref2)

Remaining priority groups were based on age: ≥85 years (priority group 3), 80-84 years (priority group 7), 75-79 years (priority group 8), 65-74 years (priority group 9), 16-64 years (priority group 10), 12-15 years (priority group 11), and 5-11 years (priority group 12). Individuals ≥85 years of age were called for vaccination early in February 2021, whereas vaccination of the remaining age groups started in mid-March with vaccinations of the 80-84-year-olds and moving down through age groups. Priority group 10 was called for vaccination in five-year age bands starting with the eldest in this priority group. Due to increased transmission of SARS-CoV-2 among the youngest part of the population, the Danish Health Authority called individuals 16-50 years of age from both ends of the age band, calling the 16-19- and 45–49-year-olds simultaneously and ending with the 30-34-year-olds. By the end of July 2021, the vaccination program was extended to include 12-15-year-olds and from end November 2021 also to children aged 5-11 years.

**Implementation of priority groups for this study.**

For the purpose of this study, some of the vaccination priority groups mentioned above have been collapsed and the following groups were evaluated:

A. Nursing home residents.
B. Healthcare- and social workers.
C. Individuals at particularly high risk of COVID-19. This includes
   a. individuals ≥65 years of age living in own homes but being in need of special care
   b. individuals with weakened immune system or obesity
   c. individuals with chronic diseases such as cancer, diabetes, chronic lung disease, kidney diseases, bowel- and liver diseases, and cardiovascular diseases.
D. Individuals who are 65 years or older and not included in any other categories.
E. Individuals who are younger than 65 years and not included in any other categories.

A graphic illustration of the Danish COVID-19 vaccine rollout and the implementation hereof in this study is presented in **Supplementary Figure 1**.

Ref1: <https://www.sst.dk/-/media/Udgivelser/2020/Corona/Vaccination/Brev-til-regioner-og-PLO-Visitation-af-personer-i-saerligt-oeget-risiko.ashx?la=da&hash=15CF482ED29FD43FD35619A7B05C55B0CE1274C3>

Ref2: <https://www.sst.dk/-/media/Udgivelser/2020/Corona/Oeget-risiko/Pjece-Personer-i-oeget-risiko---Fagligt-grundlag.ashx?la=da&hash=E7CCE677FACE1651369B2B98074721FE11B35CF9>

**Supplementary figures captions.**

**Supplementary Figure 1.** A graphic illustration of the Danish COVID-19 vaccine rollout and the implementation hereof.

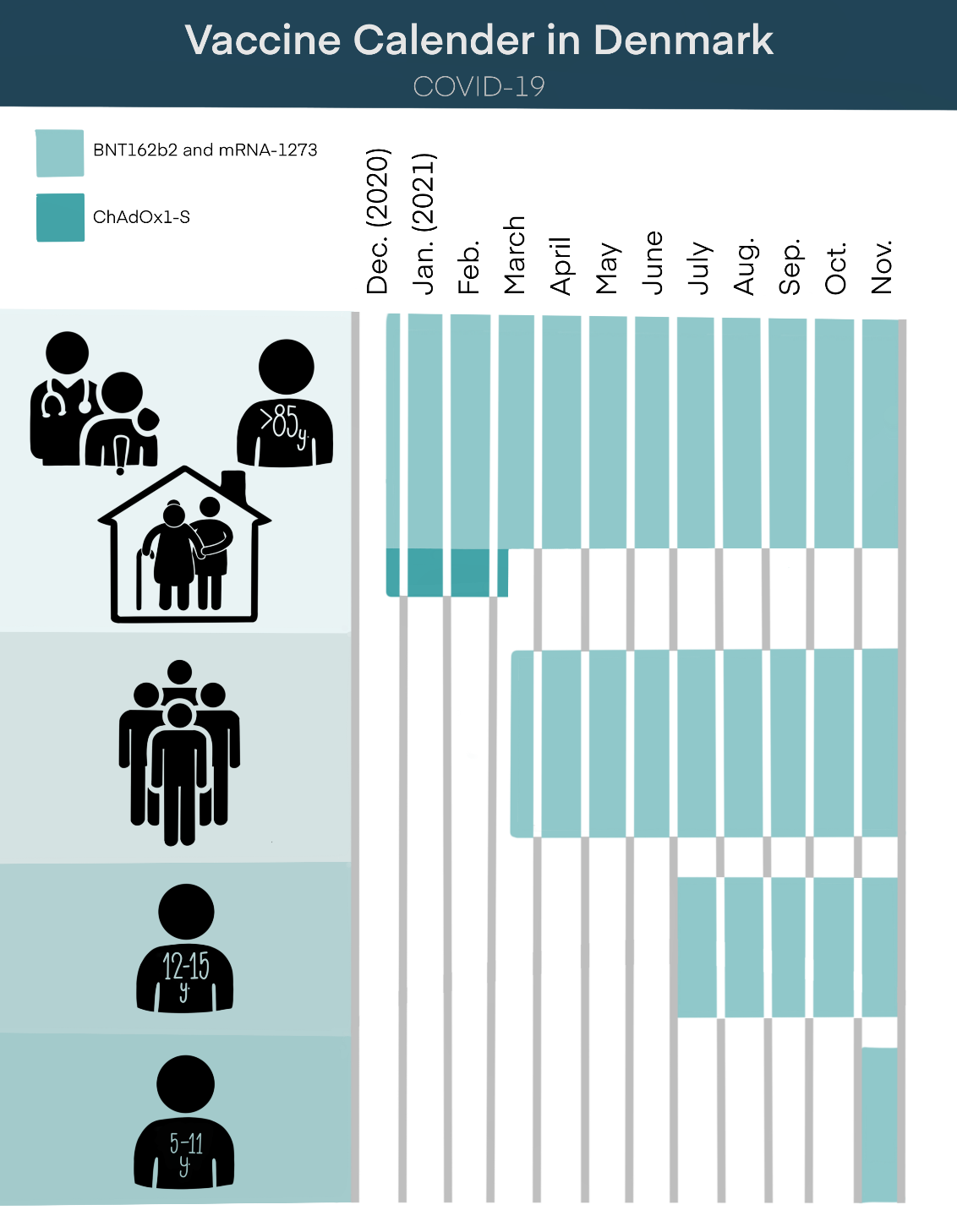

**Supplementary Figure 2.** The cumulative incidence proportion i.e., the cumulative proportion of individuals who had received their first dose of COVID-19 vaccine from December 27^th^, 2020, to January 29^th^, 2022, stratified by age groups. The denominator is the total count of Danish residents within the specific age group on January 1^st^, 2022.

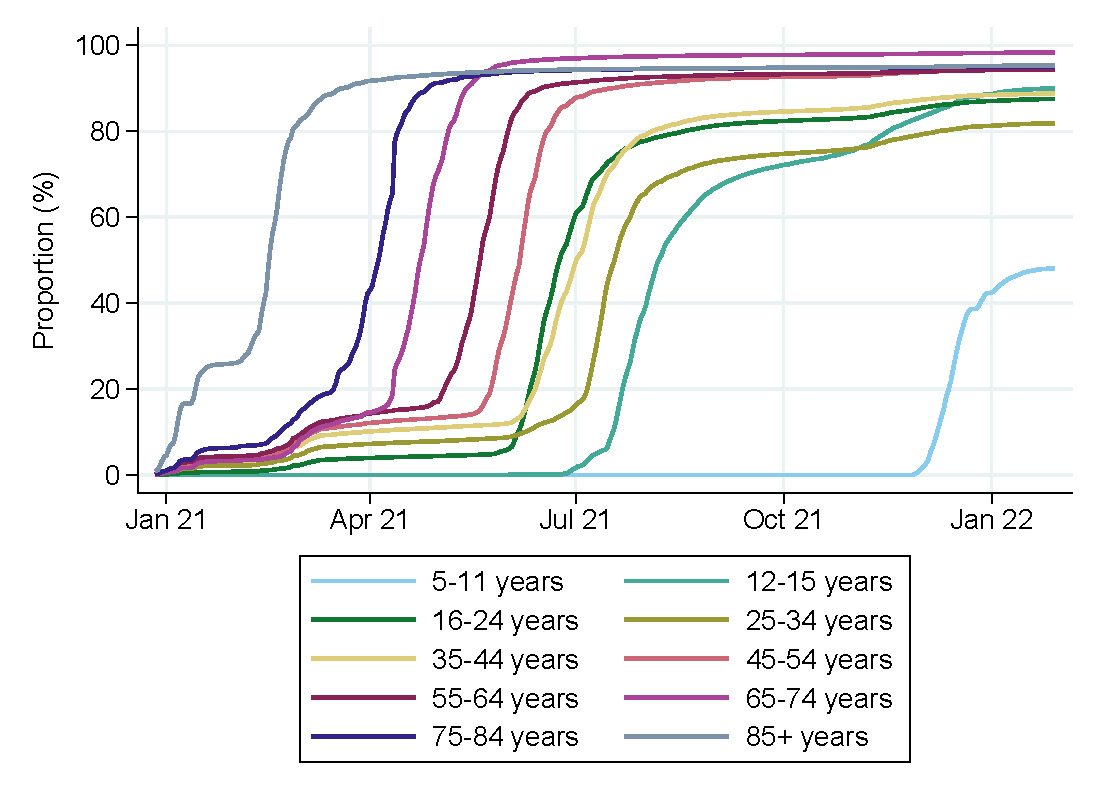

**Supplementary Figure 3.** Characteristics of individuals receiving their first dose of a COVID-19 vaccine, stratified by type of vaccine.

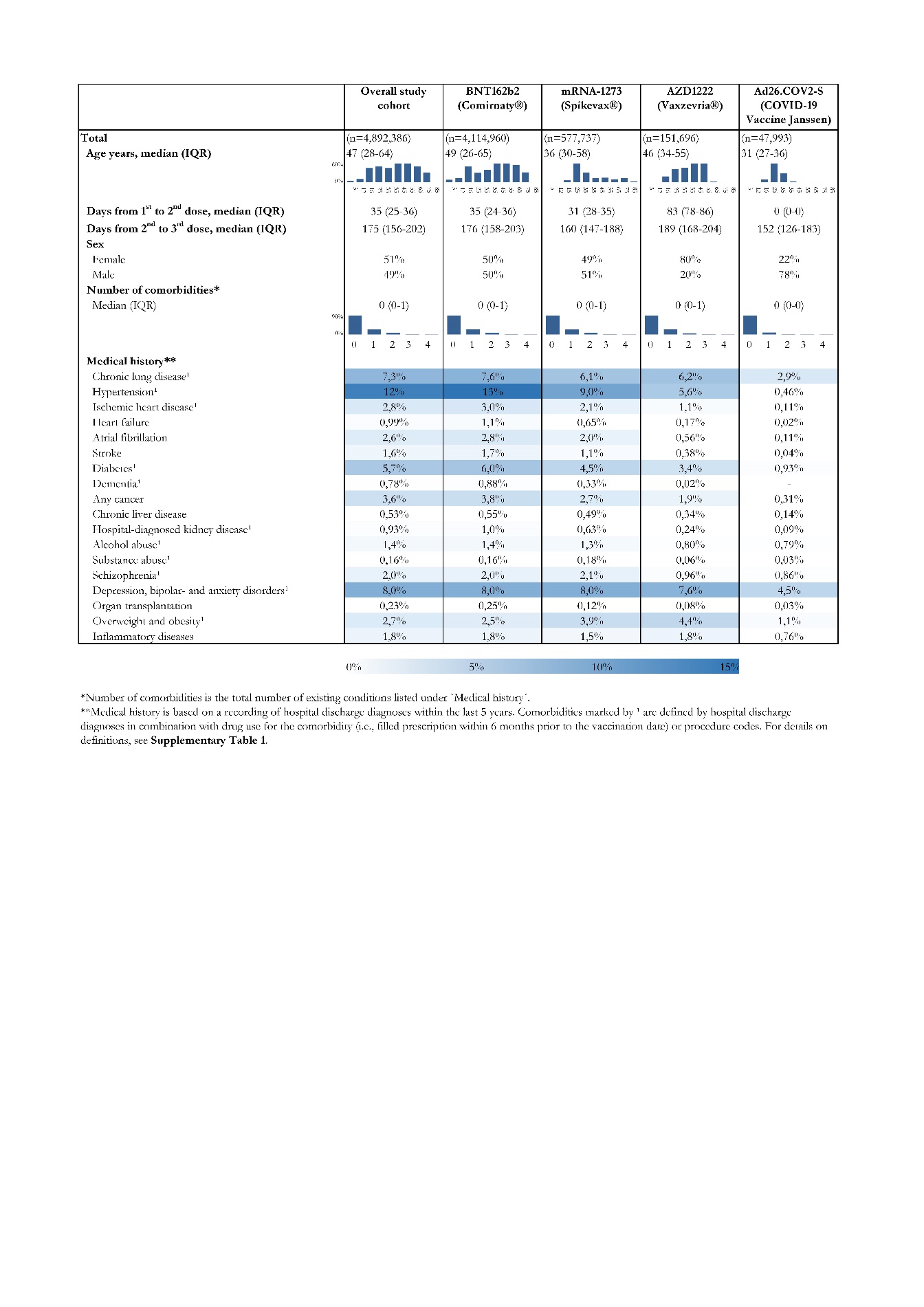

**Supplementary Figure 4.** The number of individuals who received the first dose of a COVID-19 vaccine each day from December 27^th^, 2020, to January 29^th^, 2022, stratified by type of vaccine.

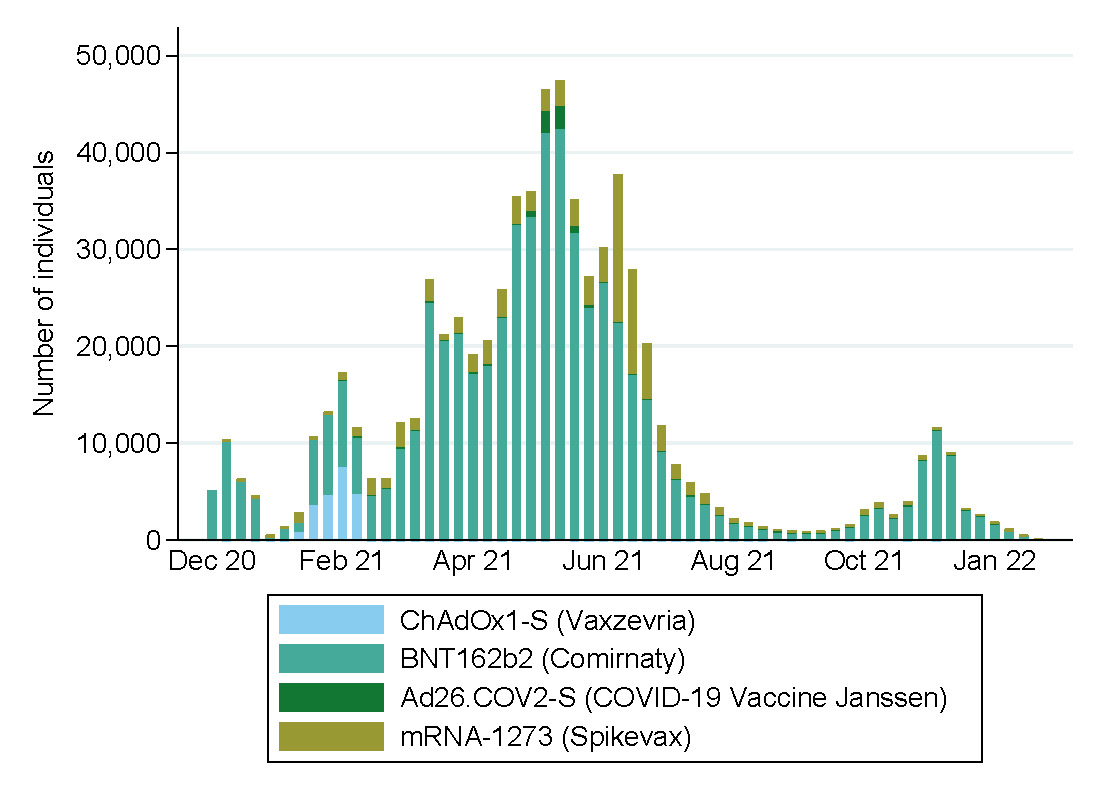

**Supplementary Tables**

**Supplementary Table 1.** ICD-10- and ATC-codes used to define comorbidity.

| Medical history^1^ | Coding system | Codes |
| --- | --- | --- |
| Chronic lung disease | ICD-10 | J41-J47 E84 |
|  | ATC | R03AK R03AL R03BA R03AC12 R03AC13 R03AC18 R03AC19 R03CC12 R03BB04 R03BB05 R03BB06 R03BB07 |
| Hypertension | ICD-10 | I10-I13 I15 |
|  | ATC | C03A, C07, C08, C09^2^ |
| Ischemic heart disease | ICD-10 | I20 I21 I22 I23 I24 I25 |
|  | ATC | B01AC24 C01DA N02BA |
| Heart failure | ICD-10 | I099A I110 I130 I132 I50 |
| Atrial fibrillation | ICD-10 | I48 |
| Stroke | ICD-10 | I60 I61 I62 I63 I64 I69 |
| Diabetes | ICD-10 | E10 E11 E13 E14 G632 H360 N083 O24 excluding O244 |
|  | ATC | A10 |
| Dementia | ICD-10 | B220A F00-F03 F1073 F1173 F1273 F1373 F1473 F1573 F1673 F1873 F1973 G30 G31 |
|  | ATC | N06D |
| Any cancer | ICD-10 | C00-C97, excluding C44 |
| Chronic liver disease | ICD-10 | B150 B160 B162 B18 B190 I85 K700-K704 K709 K71-K74 K760 K766 |
| Hospital-diagnosed kidney disease | ICD-10  Procedure codes | E102 E112 E142 I12 I13 N00-N05 N07 N08 N11 N14 N18 N19 Z992 Z49  BJFZ BJFD |
| Alcohol abuse | ICD-10 | E244 E529A F10 G312A G312B G312C G312D G312E G405B G621 G721 I426 K292 K70 K852 K860 O354 P043 T519 Z502 Z714 Z721 |
|  | ATC | N07BB |
| Substance abuse | ICD-10  ATC | F11-F19  N07BC |
| Schizophrenia | ICD-10  ATC | F20 F25  N05A |
| Depression, bipolar, and anxiety disorders | ICD-10  ATC | F30-33 F40 F41  N06A |
| Organ transplantation^3^ | ICD-10 | Z94 |
| Overweight and obesity | ICD-10  ATC | E66  A08 |
| Inflammatory diseases | ICD-10 | K50 K51 L40 M05 M06 M074 M075 M091 |

^1^Medical history is based on a recording of hospital discharge diagnoses within the last 5 years, with or without combination with drug redemption data and procedure codes.

^2^Included if patient redeemed a prescription for two different drug classes.

^3^ Medical history is based on an ever-recording of hospital discharge diagnoses.

**Supplementary Table 2.** The proportion of vaccine recipients who also receives the second and third dose, specified by age groups and vaccination priority groups.

|  | **Total number of vaccine recipients** | **Uptake of second dose** | **Uptake of third dose** |
| --- | --- | --- | --- |
| **Total** | n=4,892,386 | n=4,785,191 (98%) | n=3,563,498 (73%) |
| **Age groups** |  |  |  |
| 5-11 | 186,216 | 75% | (n<5) |
| 12-15 | 233,314 | 95% | 0.28% |
| 16-24 | 570,883 | 98% | 53% |
| 25-34 | 646,675 | 98% | 59% |
| 35-44 | 602,390 | 99% | 72% |
| 45-54 | 730,392 | 99% | 88% |
| 55-64 | 730,823 | 100% | 93% |
| 65-74 | 619,262 | 100% | 96% |
| 75-84 | 435,241 | 99% | 95% |
| 85+ | 137,190 | 98% | 87% |
| **Vaccination priority groups** |  |  |  |
| Nursing home residents | 56,242 | 97% | 78% |
| Healthcare and social workers | 300,494 | 100% | 94% |
| Individuals at particularly high risk | 285,541 | 99% | 90% |
| Others ≥65 years | 923,867 | 100% | 96% |
| Others <65 years | 3,326,242 | 97% | 63% |

**Supplementary Table 3.** Recipients of homologous and heterologous COVID-19 vaccine schedules in Denmark from December 27^th^, 2020, to January 29^th^, 2022. A) First- vs. second dose; B) Second- vs. third dose.

| A) | |  | | | |
| --- | --- | --- | --- | --- | --- |
|  | | Type of vaccine and number of recipients,  second dose (n=4,785,191) | | | |
|  | | AZD1222 (Vaxzevria®) | BNT162b2 (Comirnaty®) | Ad26.COV2-S (COVID-19 Vaccine Janssen) | mRNA-1273 (Spikevax®) |
| Type of vaccine and number of recipients, first dose  (n=4,892,386) | AZD1222 (Vaxzevria®) | 1599 (1.1%) | 96,721 (64%) | - | 52,365 (35%) |
|  | BNT162b2 (Comirnaty®) | 9 (0%) | 4,016,885 (100%) | - | 162 (0%) |
|  | Ad26.COV2-S (COVID-19 Vaccine Janssen) | N/A* | N/A* | 47,993* | N/A* |
|  | mRNA-1273 (Spikevax®) | - | 311 (0.05%) | - | 569,146 (100%) |
| B) | | | | | |
|  | | Type of vaccine and number of recipients,  third dose (n=3,563,498) | | | |
|  | | AZD1222 (Vaxzevria®) | BNT162b2 (Comirnaty®) | Ad26.COV2-S (COVID-19 Vaccine Janssen) | mRNA-1273 (Spikevax®) |
| Type of vaccine and number of recipients, second dose  (n=4,785,191) | AZD1222 (Vaxzevria®) | 6 (0.48%) | 1,173 (94%) | - | 69 (5.5%) |
|  | BNT162b2 (Comirnaty®) | 270 (0.01%) | 3,067,104 (100%) | - | 736 (0.02%) |
|  | Ad26.COV2-S (COVID-19 Vaccine Janssen) | - | 4,665 (11%) | 28 (0.07%) | 36,633 (89%) |
|  | mRNA-1273 (Spikevax®) | 122 (0.03%) | 1,290 (0.28%) | - | 451,402 (100%) |

*One dose of AD26.COV2-S vaccine (COVID-19 Vaccine Janssen) contains both the first and the second dose.
